# Determinants of Late Initiation for Antenatal Care Follow up Among Pregnant Mothers Attending Public Health Centers at Jimma Town, South West Ethiopia, 2021: Unmatched Case–Control Study

**DOI:** 10.1101/2022.01.20.22269582

**Authors:** Mohammed Jihad, Kifle Woldemichael, Yenealem Gezehagn

## Abstract

**Background:** World Health Organization recommends that, a woman without complications should have at least four antenatal care visits, the first of which should take place during the first trimester (not > 16 weeks). But, high proportion of pregnant women in most of developing countries attended first ANC at a late time. In Ethiopia, magnitude of late ANC follow up initiation is 64% and in study area 68.7% however determinants of the problem were not identified particularly in the study area.

**Objective:** To identify determinants of late antenatal care initiation among pregnant women attending antenatal care in public health centers of Jimma town, south west Ethiopia 2021.

**Methods:** Facility based unmatched case control study was conducted from May 1-30. The cases (n=177) were mothers who attended their first antenatal care visit at >= 16 weeks of gestational age and the controls (n=177) were mothers who attended their first antenatal care visit at <16 weeks of gestational age during the data collection period. Data was collected through face to face interview using pre tested structured questionnaire. Descriptive and summary statistics was done. Bivariable and multivariable logistic regression analysis was done using SPSS version 21.0 to identify candidate variables at P-value <0.25 and determinants of late ANC initiation at P-value <0.05 respectively.

**Results:** The current study identified that, being age > 25 years (AOR= 2.623, 95% CI 1.059 - 6.496), poor knowledge about ANC (AOR= 3.856, 95% CI 1.266-11.750), lack of history of abortion (AOR= 3.326, 95% CI 1.082-10.224), place of previous delivery (AOR=3.926, 95%CI 1.023-15.063), recognizing pregnancy through missed period(AOR= 3.631, 95% CI 1.520-8.674), pregnancy related pain(AOR= 3.499, 95% CI 1.423 - 8.603) and lack of family support (AOR=2.647, 95% CI 1.092-6.415) were determinants of late ANC initiation.

**Conclusion:** Being age > 25 years, poor knowledge about ANC, lack of history of abortion, place of previous delivery, recognizing pregnancy through missed period, pregnancy related pain, and lack of family support during ANC booking were determinants of late ANC initiation.Health facilities and respective health care providers should provide health education at facility level regularly with due attention to late ANC initiation.

## Introduction

Late ANC initiation is the first ANC visit at >= 16 weeks of gestational age for the current pregnancy. Under normal condition, World Health Organization recommends that a woman without complications should have at least four antenatal care visits, the first of which should take place during the first trimester (not > 16 weeks) (1).There are a number of risk factors associated with late antenatal booking and these may have an impact on maternal and foetal wellbeing, resulting in increased mortality and morbidity (1–4).

Worldwide there is a big discrepancy in the prevalence of late ANC follow up among pregnant mothers, ranging from 27.5 to 88% in developed and developing countries respectively. Delayed initiation of antenatal follow up is the predominant problems throughout Sub Saharan African (SSA) countries including Ethiopia resulting in failure of accomplishment of the World Health Organization recommendation (5,6).

According to the systematic review and meta-analysis conducted on delayed initiation of ANC and associated factors in Ethiopia, the pooled estimate of the magnitude of delayed initiation of ANC in Ethiopia was 64%(7). High proportion of the pregnant women (60.5%) initiated antenatal care after four months of gestation (8–10). Mini EDHS 2014 revealed that, only 18% of pregnant women initiated early for first ANC services (11). EDHS 2016 also showed that, 42% of first ANC attendants were initiated lately (12). Study conducted in Jimma town, Ethiopia showed that 68.7% of participants initiated for their first ANC lately(13).

Late ANC registration and inadequate ANC attendance predispose pregnant women to increased risk of maternal morbidity and mortality and poor pregnancy outcome. Women who book late for ANC lose the opportunity to benefit from early detection and effective treatment of some disease conditions that may impair their health and that of their babies(14). Low birth weight, low Apgar scores as well as the incidence of maternal anaemia and pregnancy induced hypertension (PIH) were found to be related to late ANC booking(15).

Adverse pregnancy outcomes can be minimised or avoided altogether if antenatal care is received early in the pregnancy and continued through delivery. But every day in 2017, about 808 women died due to complications of pregnancy and child birth. MMR in the world’s least developed countries (LDCs) is high, estimated at 415 maternal deaths per 100 000 live births, which is more than 40 times higher than MMR in Europe, and almost 60 times higher than in Australia and New Zealand.Almost all of these deaths occurred in low-resource settings, and most could have been prevented. The primary causes of death are haemorrhage, hypertension, infections, and indirect causes, mostly due to interaction between pre-existing medical conditions and pregnancy(16,17).

Many studies reported that, late initiation of ANC was highly significant among less educated mother, unplanned pregnancy, age >26 years of mothers, unemployed women, husband’s education, women’s autonomy, knowledge on ANC, partner involvement, past pregnancy complication history, parity, anxiety of being pregnant, monthly income, place of residence, revealing pregnancy, means of checking pregnancy, late recognition of pregnancy, distance of health facility, place of previous delivery and not having experience of ANC(7,8,18–22).

In Ethiopia many studies indicated that, ANC utilization during the first trimester is low and most pregnant women who attend ANC come too late for their first ANC visit ranges from 40 to 60%. Due to this reason, only four in 10 women (43%) had four or more ANC visits for their most recent live birth.According to Ethiopian EDHS report; the ANC service coverage was 41%, among those 23% of them initiated late for ANC (3,11).

Despite having made considerable progress on an international level in terms of increasing the accessibility and use of antenatal care, the first consultation usually occurs at a late stage of pregnancy(23). Annual report of Jimma town 2019/20 from DHIS2 also showed that high proportion 64% of first ANC attendants were initiated lately.

In various parts of Ethiopia, numerous studies have been undertaken and written. However, the majority of them were designed to be cross-sectional, making it impossible to determine a causal association between the outcome and exposure variables. In addition, the determinants of late initiation for ANC are not the same across different cultures, socio economic status and access to institution within a society. As a result, the aim of this research is to find potential determinants of late ANC initiation among pregnant women attending ANC in public health centers in Jimma town, south west Ethiopia, in the year 2021.

## Methods and Materials

### Study area, designand period

This study was conducted in Oromia Regional State, Jimma Town, which is 346 km far from Addis Ababa to the southwest part of the country. Based on the 2007 national census, the estimated population of the Jimma town in 2020/21 is 220,609. Of these, 112,511 (51%) are females and 7655 Pregnant mothers. The town has 17 kebeles; 4 rural and 13 urban kebeles. There are 4 health centers, 2 public Hospitals and 3 private Hospitals in Jimma town. The data collection period was from May 1-30, 2021.

Facilitybased unmatched casecontrol studydesign wasused

### Population

Source population were all pregnant women attending ANC in Jimma town public health centers during the study period. Study population for cases were all eligible pregnant women who came for first antenatal care services at >=16 weeks of gestational age and study population for controls were all eligible pregnant women who came for first antenatal care services at <16 weeks of gestational age in Jimma town public health centers during the study period.

### Samplesize and sampling procedure

Sample size was calculated by Epi info Version 7.0.8.3 using double population proportion formula by considering proportion of mothers who had unplanned pregnancy among controls was 8.2% (main predictor variable with AOR = 2.73 and P-value <0.000) from similar study conducted in Addis Ababa, Ethiopia (24). In addition 95% CI, 80% power and 1:1 control to case ratio. Accordingly by adding 10% none response rate, the final sample size were 177 cases and 177 controls. There are four public health centers in Jimma town and the total sample size was proportionately allocated across all health centers according to their average three month’s cases load. (Figure 1)

**Figure 1:**
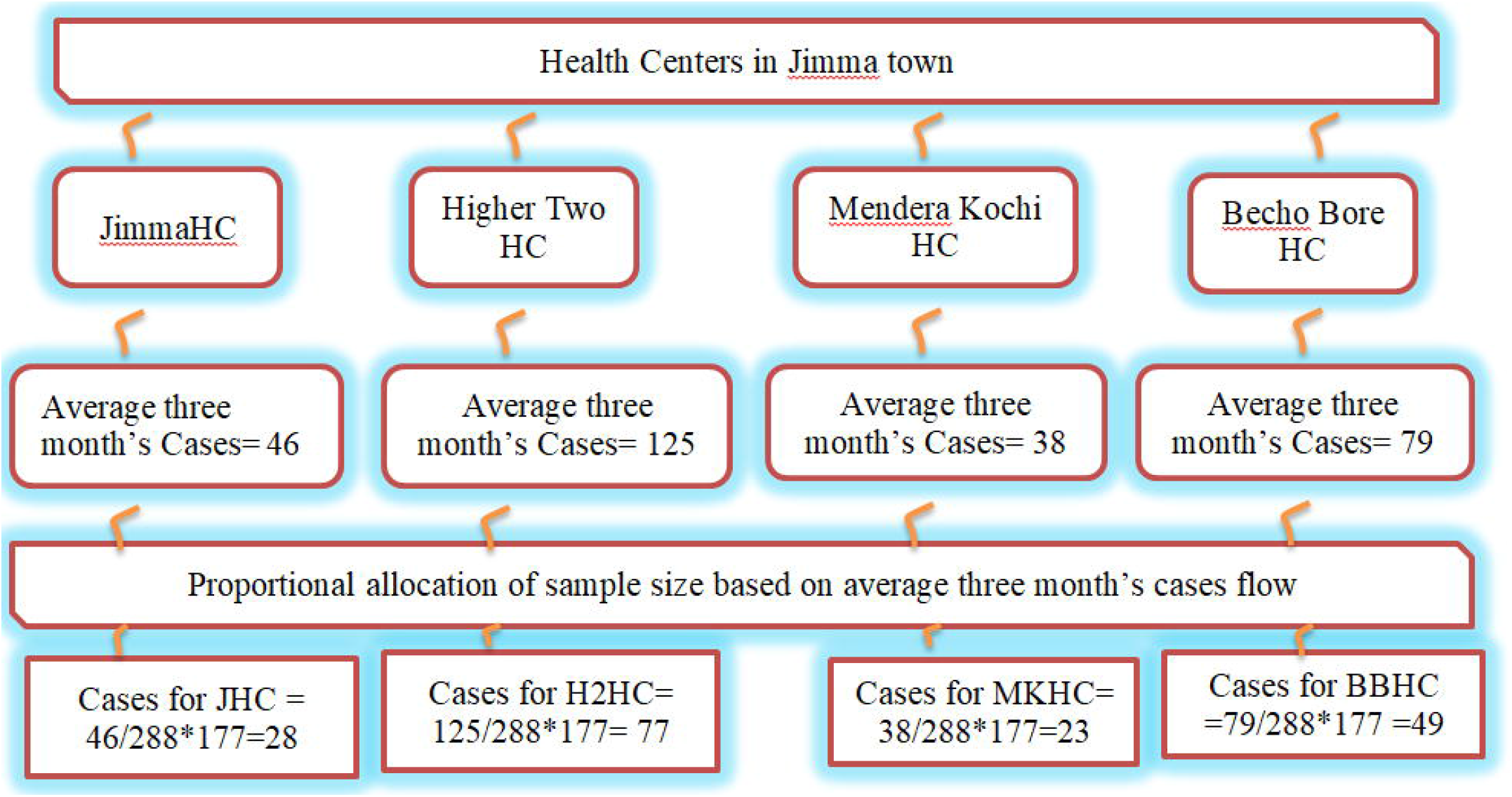
Proportional allocation of sample size among health centers in Jimma Town 2021.

#### Selection of cases

for a women’s who came for First ANC gestational age of the pregnancy was determined using LMP. Pregnant women’s with gestational age of >=16 weeks (cases) was identified and selected using systematic random sampling technique. The random start of the case selection was 4 and the interval was 2.

The interval was determined by average three month’s cases flow divided by the sample size allocated to each facility, which is equal to two. The first mother was identified using simple random sampling and the rest were selected every interval until the allocated sample size was reached.

#### Selection of controls

Control selection was based on case selection; for each case selected, the immediate pregnant women with a gestational age < 16 weeks (control) was identified and selected as a control.

### Measurement of Variables

#### Socio-economic status

It was measured by using the information from the asset based wealth index & principal component analysis (PCA) on SPSS was used for its analysis; in that the family’s wealth was grouped in quintiles (from lowest to highest). The quintiles was Q1= Poor socio-economic level, Q2=Medium socio-economic level, and Q3= Wealthiest.

#### Knowledge

Measured based on five multiple choice questions and for correct response, 1 score was given, for incorrect or don’t know the answer, 0 was given. The cumulative score ranges from 0 to 5. Finally, by using modified Bloom’s cut off point, categorization was made as, 80-100% good knowledge, 50-79% moderate and <50% poor knowledge.

#### Service quality

Measured by using Likert’s scale based on satisfaction level of the mothers on health service delivery. Four questions were rated by mothers and score was given starting from 5-1, the cumulative score ranges from 4-20 then using modified Bloom’s cut off point; 80-100% was considered as good quality, 50-79% moderate quality and <50% indicates poor quality.

### Data collection tools and procedures

Structured questionnaire that has different sections was developed from different literatures. The questionnaire was first prepared in English and translated into Afan Oromo and back translated to English to cheek consistency of the language. Data collectors were eight diploma midwives and one BSc midwife supervisor has followed day to day activities. Two days training was provided on objectives of the study, subject identification and contents of questionnaire for data collectors, supervisor and data clerks. Data was collected through face to face interview at waiting area following the ANC consultation at ANC unit. Respondents were informed about the purpose of the study, privacy, confidentiality and informationgiven byeach respondent was kept in a safe boxand namewas not recorded. Data was checked by the supervisor at the end of each day during the data collection period, whether the questionnaires were complete and consistent. After data collection, questionnaires were sorted into two groups and numbered as 1 and 2 controls and cases respectively.

### Data analysis procedure

Data was entered into EPI-data version 4.6 and exported to SPSS version 21.0 for analysis. Descriptive statistics was used to summarize the data according to the variable type. Bivariable and multivariable logistic regression were done. Each variable with the outcome of interest at p < 0.25 in the bivariable analysis wascandidate for multivariable analysis. Multi-collinearity was checked for all candidate variables using VIF (Variance Inflator Factor) at the cut of point 10 and Model goodness of fit using the Hosmer & Lemeshow test at p value > 0.05 was done. In the multivariable analysis, P < 0.05 declared statistically significant. AOR with its respective 95%CI was calculated to measure the association between independent variables and a dependent variable.

### Ethical Consideration

Ethical clearance was obtained from ethical review board (IRB) of Jimma University. Based on the ethical clearance, an official letter was obtained from the Institute of Health, Faculty of Public Health, Department of Epidemiology. An official letter was submitted to the Jimma town health office and health centers. After explaining the purpose of the study, permission was obtained from the head of Jimma town health office and managers of health centers. Respondents were informed about the purpose of the study, the importance of their participation, and the right to withdraw at any time if they want and about privacy and confidentiality of the information given by each respondent kept properly and that name was not recorded. Written consent was obtained from each respondent.

## Results

### Socio demographic characters

In the current study 177 cases and 177 controls participated with a response rate of 100%. About 151(85.3%) of cases and 162 (91.5%) controls were urban residents. The distribution of religion types among cases and controls was nearly the same. Regarding age of the mothers, 96(54.2%) and 43(24.3%) of cases and of controls were age >25 years respectively.

Almost all cases 168 (94.9%) and controls 167 (94.4%) were married. One hundred twenty eight cases (72.3%) and 111(62.7%) controls were unemployed. Regarding to husband occupation, 110(64%) of cases and 134 (76.6%) of controls were employed. Concerning socio economic status, 63(35.6%) of cases and 55(31.1%) controls were in the category of poorest socio-economic status. While 57(32.2%) of cases and 61(34.5%) controls were wealthiest (Table 1).

**Table 1:** Socio demographic characteristics of mothers for the research conducted on determinants of late ANC initiation, in Jimma town, South West Ethiopia, 2021.

| Variables | Categories | Case n= 177 | Control n =177 | P-Value | COR(95% CI) |
| --- | --- | --- | --- | --- | --- |
| Residence | Urban | 151(85.3%) | 162(91.5%) | 1 |  |
|  | Rural | 26(14.7%) | 15(8.5%) | .271 | 0.538(0.274-1.054) |
| Religion | Muslim | 126(71.2%) | 128(72.3%) | 1 |  |
|  | Orthodox | 38(21.5%) | 37(20.9%) | .872 | 0.958(0.573-1.604) |
|  | Protestant | 11(6.2%) | 12(6.8%) | .870 | 1.074(0.457-2.523) |
|  | Other | 2(1.1%) | 0(0%) | .999 | .000 |
| Age of mother | >25 | 96(54.2%) | 43(24.3%) | .000 | 3.693(2.347-5.811) |
|  | <=25 | 81(45.8%) | 134(75.7%) | 1 |  |
| Family size | >3 | 76(42.9%) | 39(22%) | .000 | 0.376(0.236-.597) |
|  | <=3 | 101(57.1%) | 138(78%) | 1 |  |
| Marital status | Single | 7(4.0%) | 7(4.0%) | .991 | 1.006(0.345-2.931) |
|  | Divorced/widowed | 2(1.1%) | 3(1.7%) | .655 | 1.509(0.249-9.147) |
|  | Married | 168(94.9%) | 167(94.4%) | 1 |  |
| Occupational status of mother | Unemployed | 128(72.3%) | 111(62.7%) | .054 | 1.553(0.99-2.432) |
|  | Employed | 49(27.7%) | 66(37.3%) | 1 |  |
| Occupational status of husband | Unemployed | 62(36%) | 41(23.4%) | .011 | 1.842(1.153-2.942) |
|  | Employed | 110(64%) | 134(76.6%) | 1 |  |
| Educational status of mother | Can't read and write | 44(24.9%) | 16(9%) | .000 | 4.583(2.284-9.197) |
|  | primary (1-8) | 94(53.1%) | 96(54.2%) | .002 | 2.809(1.482-5.321) |
|  | Secondary (9-12) and above | 39(22%) | 65(36.7%) | 1 |  |
| Educational status of husband | Can't read and write | 26(15.1%) | 15(8.6%) | .002 | 3.026(1.485-6.164) |
|  | primary (1-8) | 87(50.6%) | 57(32.6%) | .728 | 1.136(0.554-2.328) |
|  | secondary (9-12) and above | 59(34.3%) | 103(58.9%) | 1 |  |
| Socio economic status | Poorest | 56(31.6%) | 68(38.4%) | .322 | 1.495(0.898-2.486) |
|  | Moderate | 57(32.2%) | 57(32.2%) | .432 | 1.231(0.733-2.067) |
|  | Wealthiest | 64(36.2%) | 52(29.4%) | 1 |  |

### Knowledge status of respondents about ANC

Seventy-seven (43.5%) of cases and 37(20.9%) controls have poor knowledge about ANC. Proportion of moderate knowledge among cases and controls was 67(37.9%) and 76 (42.9%) respectively. Thirty three (18.6%) of cases and 64(36.2%) of controls have good knowledge about ANC (Figure 2).

**Figure 2:**
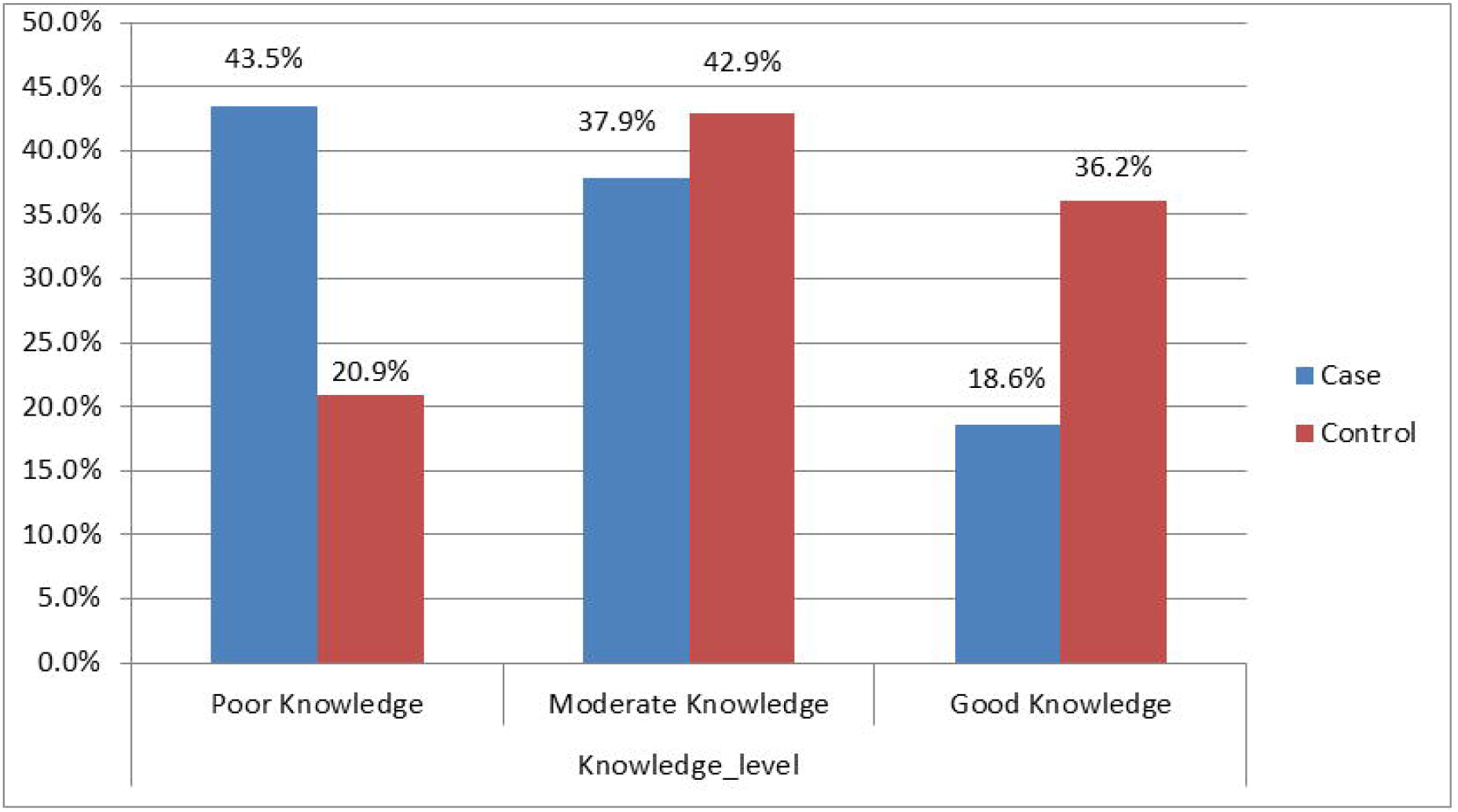
Knowledge status of mothers among case and control groups, Jimma Town, South West Ethiopia, 2021.

### Maternal and obstetric characteristics of respondents

Proportion of multigravida among cases and controls was 112(63.3%) and 81(45.8%) respectively. Sixty five (36.7%) of cases and 96(54.2%) controls were primi-gravida. Mothers were asked if they have had history of abortion and 18(16.1%) of cases and 28(34.6%) controls reported history of abortion. Concerning place of delivery for the last previous pregnancy, 25(23.6%) of cases and 7(10%) of controls gave birth at their home. Three fourth 133(75.1%) of cases and 68(38.4%) of controls recognized their pregnancy through missed period. Regarding torevealing being pregnantto others, 137(77.4%) of cases and 169(95.5%) of controls revealed their pregnancy (Table 2).

**Table 2:**
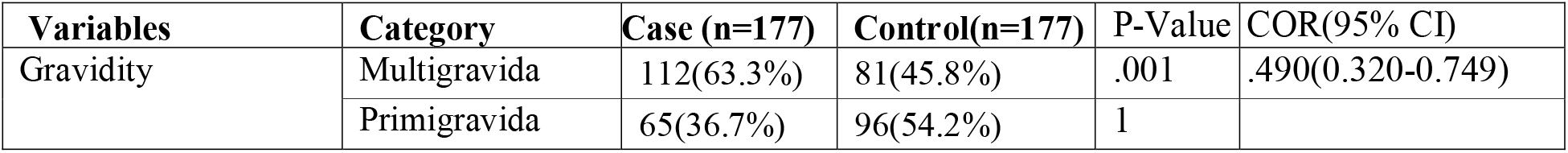
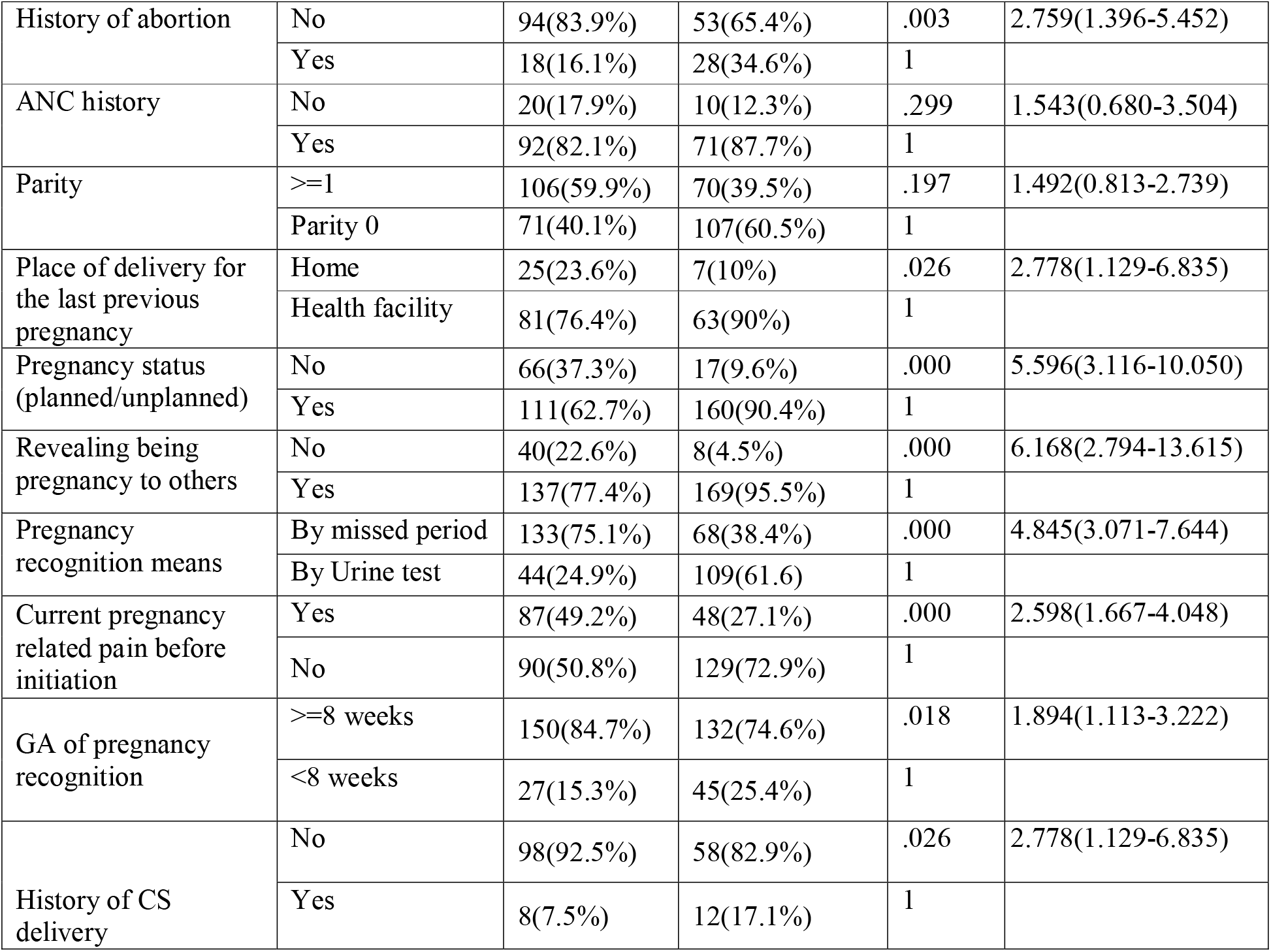
Bivariable Analysis of Maternal and Obstetric factorsassociated with late ANC initiation, in Jimma town, South West Ethiopia, 2021.

| Variables | Category | Case (n=177) | Control(n=177) | P-Value | COR(95% CI) |
| --- | --- | --- | --- | --- | --- |
| Gravidity | Multigravida | 112(63.3%) | 81(45.8%) | .001 | .490(0.320-0.749) |
|  | Primigravida | 65(36.7%) | 96(54.2%) | 1 |  |
| History of abortion | No | 94(83.9%) | 53(65.4%) | .003 | 2.759(1.396-5.452) |
|  | Yes | 18(16.1%) | 28(34.6%) | 1 |  |
| ANC history | No | 20(17.9%) | 10(12.3%) | .299 | 1.543(0.680-3.504) |
|  | Yes | 92(82.1%) | 71(87.7%) | 1 |  |
| Parity | >=1 | 106(59.9%) | 70(39.5%) | .197 | 1.492(0.813-2.739) |
|  | Parity 0 | 71(40.1%) | 107(60.5%) | 1 |  |
| Place of delivery for the last previous pregnancy | Home | 25(23.6%) | 7(10%) | .026 | 2.778(1.129-6.835) |
|  | Health facility | 81(76.4%) | 63(90%) | 1 |  |
| Pregnancy status (planned/unplanned) | No | 66(37.3%) | 17(9.6%) | .000 | 5.596(3.116-10.050) |
|  | Yes | 111(62.7%) | 160(90.4%) | 1 |  |
| Revealing being pregnancy to others | No | 40(22.6%) | 8(4.5%) | .000 | 6.168(2.794-13.615) |
|  | Yes | 137(77.4%) | 169(95.5%) | 1 |  |
| Pregnancy recognition means | By missed period | 133(75.1%) | 68(38.4%) | .000 | 4.845(3.071-7.644) |
|  | By Urine test | 44(24.9%) | 109(61.6) | 1 |  |
| Current pregnancy related pain before initiation | Yes | 87(49.2%) | 48(27.1%) | .000 | 2.598(1.667-4.048) |
|  | No | 90(50.8%) | 129(72.9%) | 1 |  |
| GA of pregnancy recognition | >=8 weeks | 150(84.7%) | 132(74.6%) | .018 | 1.894(1.113-3.222) |
|  | <8 weeks | 27(15.3%) | 45(25.4%) | 1 |  |
| History of CS delivery | No | 98(92.5%) | 58(82.9%) | .026 | 2.778(1.129-6.835) |
|  | Yes | 8(7.5%) | 12(17.1%) | 1 |  |

### Health service related factors

Regarding service quality, 82(46.3%) of cases and 84(47.5%) controls rated service quality as moderate. Fifty two (29.4%) and 29(16.4%) of cases and controls rated service quality as poor respectively. Concerning distance from health facility, 118(66.7%) of cases and 130(73.4%) of controls were near to health facility (Table 3).

**Table 3:**
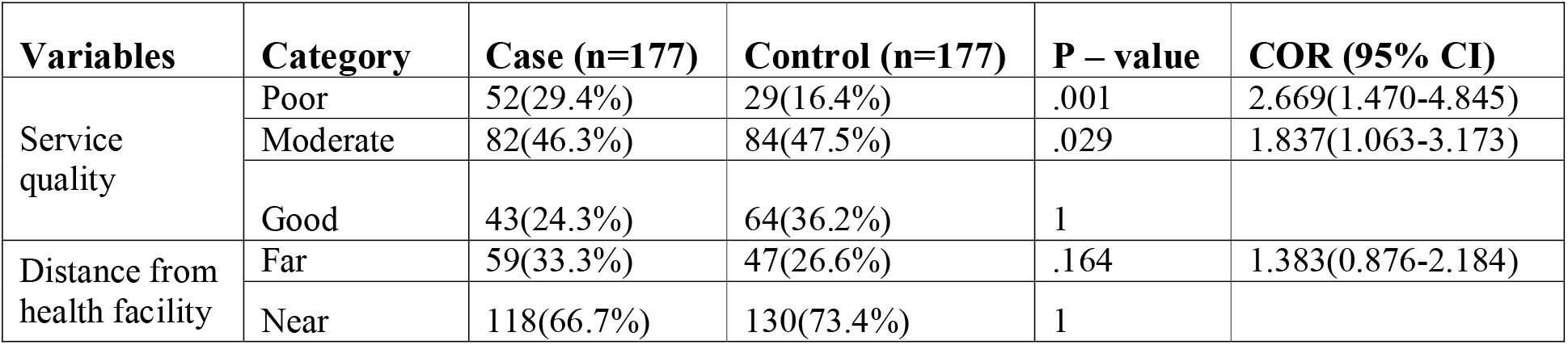
Bivariable Analysis of Health service related factorsassociated with late ANC initiation, in Jimma town, South West Ethiopia, 2021.

### Socio-cultural characteristics

One hundred twenty-six (71.2%) of cases and 158 (89.3%) of controls made decisions to book for ANC follow up in consultation with their husbands. Regarding family support, 118(66.7%) of cases and 55(31.1%) controls came to health facility alone during ANC initiation (Table 4).

**Table 4:** Socio-cultural characteristics of respondents, Jimma Town, South West Ethiopia 2021.

| Variables | Categories | Case (n= 177) | Control (n= 177) | P-Value | COR (95%CI) |
| --- | --- | --- | --- | --- | --- |
| Decision making to book for ANC | My Self | 45(25.4%) | 16(9%) | .000 | 3.527(1.904-6.534) |
|  | with Other relative | 6(3.4%) | 3(1.7%) | .656 | 1.406(0.314-6.294) |
|  | with my husband | 126(71.2%) | 158(89.3%) | 1 |  |
| Family support | Alone | 118(66.7%) | 55(31.1%) | .000 | 5.185(3.259-8.248) |
|  | with Other relative | 11(6.2%) | 6(3.4%) | .768 | 1.170(0.412-3.327) |
|  | with my husband | 48(27.1%) | 116(65.5%) | 1 |  |

### Determinants of late initiation of ANC follow-up

Mothers aged >25 years were 2.623 times more likely to initiate ANC follow-up late as compared to their counterparts (AOR= 2.623, 95% CI 1.059 - 6.496). Pregnant mothers who have poor knowledge about ANC were 3.856 times more likely to book late for ANC as compared to those who have had good knowledge (AOR= 3.856, 95% CI 1.266- 11.750).Mothers who hadn’t history of abortion were 3.326 times more likely to book late for ANC follow-up as compared to those who had an abortion history (AOR= 3.326, 95% CI 1.082 −10.224). Mothers who gave birth at their home during the last previous delivery were 3.9 times more likely to book late for ANC as compared to mothers who gave birth at health facility (AOR=3.926, 95%CI 1.023-15.063).

Pregnant mothers who recognized their pregnancy by missed period were 3.631 times more likely to book late for ANC follow-up as compared to those recognized by urine test (AOR= 3.631, 95% CI 1.520-8.674). Mothers who wait until they feel ill of pregnancy were 3.499 times more likely to book late for ANC follow-up as compared to those who initiated without feeling pain of pregnancy (AOR= 3.499, 95% CI 1.423 - 8.603). Mothers who came to health facilities alone during ANC booking were 2.647 times more likely to initiate late for ANC follow up as compared to those came with their husbands (AOR=2.647, 95% CI 1.092 - 6.415) (Table 5).

**Table 5:** Independentpredictorsof late ANC initiation, Jimma Town, South West Ethiopia 2021.

| Variables | Case(n=177) | Control(n=177) | COR (95% CI) | AOR (95% CI) |
| --- | --- | --- | --- | --- |
| **Age of mother** |  |  |  |  |
| >25 | 96(54.2%) | 43(24.3%) | 3.693 (2.347-5.811) * | 2.623(1.059-6.496) ** |
| <=25 | 81(45.8%) | 134(75.7%) | 1 |  |
| **Knowledge about ANC** |  |  |  |  |
| Poor Knowledge | 77(43.5%) | 37(20.9%) | 4.036(2.272-7.170) * | 3.856(1.266-11.750) ** |
| Moderate Knowledge | 67(37.9%) | 76(42.9%) | 2.361(1.415-3.937) * | 1.98(0.424-3.381) |
| Good Knowledge | 33(18.6%) | 64(36.2%) | 1 |  |
| **History of abortion** |  |  |  |  |
| No | 94(83.9%) | 53(65.4%) | 2.759 (1.396-5.452) * | 3.326(1.082-10.224) ** |
| Yes | 18(16.1%) | 28(34.6%) | 1 |  |
| **Place of delivery for the last previous pregnancy** |  |  |  |  |
| Home | 25(23.6%) | 7(10%) | 2.778 (1.129-6.835) * | 3.926(1.023-15.063) ** |
| Health facility | 81(76.4%) | 63(90%) | 1 |  |
| **Pregnancy recognition Means** |  |  |  |  |
| By missed period | 133(75.1%) | 68(38.4%) | 4.845 (3.071-7.664) * | 3.631(1.520-8.674) ** |
| By Urine test | 44(24.9%) | 109(61.6) | 1 |  |
| **Current pregnancy related pain before initiation** |  |  |  |  |
| Yes | 87(49.2%) | 48(27.1%) | 2.598 (1.667-4.048) * | 3.499(1.423-8.603) ** |
| No | 90(50.8%) | 129(72.9%) | 1 |  |
| **Family support** |  |  |  |  |
| Alone | 118(66.7%) | 55(31.1%) | 5.185(3.259-8.248) * | 2.647 (1.092-6.415) ** |
| with Other relative | 11(6.2%) | 6(3.4%) | 1.170(0.412-3.327) | 1.323(0.133-13.139) |
| with my husband | 48(27.1%) | 116(65.5%) | 1 |  |
| * Significant variables in bivariable analysis, ** Significant variables in Multivariable analysis |  |  |  |  |

## Discussion

The current study showed that, being age > 25 years, poor knowledge about ANC, lack of history of abortion, previous home delivery, recognizing pregnancy through missed period, current pregnancy related pain before initiation, and lack of family support during ANCbookingwere determinants of late ANC initiation.

Pregnant mothers aged >25 were 2.6 times more likely to book late for ANC follow-up as compared to their counterparts. This result is consistent with the studies done in Debremarkos, Kembata, and East Wellega, Ethiopia (5,25,26). The reason might be due to that young women have more health seeking behaviour and information about the importance of early antenatal care booking than older women.

Pregnant mothers who have had poor Knowledge about ANC were 3.38 times more likely to book late for ANC follow up as compared to those who have had good knowledge.This finding is in line with the study conducted in Tigray, Ethiopia (24). This could be explained by the fact that mothers with poor knowledge might not have awareness about the correct time for ANC booking, frequency of ANC visits, and the importance of ANC follow up both for the mother and the foetus. In the current study, pregnant mothers who had no history of abortion were 3.3 times more likely to book late for ANC follow up as compared to their counterparts. The study finding is supported by the study done in the United Arab Emirates and in Ethiopia(20,28). It could be explained by the fact that, mothers who have a history of abortion were more likely to initiate early for ANC follow up due to fear of recurrence of the event as well as they might got information about ANC follow up.

The finding showed that, mothers who gave birth at their home during the last previous delivery were 3.9 times more likely to book late for ANC follow up as compared to those who gave birth at health institution during previous delivery. This finding is in line with the study conducted in Ethiopia (21). It might be due to mothers who gave birth at the health facility during previous delivery benefitted and get information about ANC follow up and appropriate booking time. Mothers who recognized their pregnancy through missed period were 3.6 times more likely to book for ANC late as compared to mothers who recognized their pregnancy by urine test. Similar finding was also reported in studiesconducted in North Ethiopia (5,29,30). Mothers who recognize their pregnancy through urine test might be more likely to practice health care seeking and more likely to accept modern medical care follow up than those who recognized via missed period. On the other hand, mothers who recognized pregnancy by missed period were most probably after missing one or two cycles and thereafter, they might be reluctant to start ANC soon(29).

Pregnant mothers who wait until they feel ill of pregnancy to start ANC follow up were about 3.5 times more likely to book for ANC follow up late as compared to their counterparts. This finding is consistent with the study conducted in Addis Ababa, Ethiopia(31). The possible explanation could be due to the wrong perception of ANC follow up as a curative rather than preventive and waiting pregnancy related medical problem to book for ANC. Pregnant mothers who came alone to health facilities during ANC booking were 2.64 more likely to initiate ANC follow up lately as compared to those accompanied by their husbands. This finding is supported by the study done in Tanzania and Ethiopia (9,19,32). This could be due to a lack of encouragement from husbands, which may deter women from obtaining early ANC care.

The current study showed that, family size, parity, pregnancy status and revealing being pregnancy to others were not significantly associated with late ANC booking. Family size has no statistically significant relationship with late ANC booking. The result is different from the study conducted in sub-Saharan Africa and Cameroon (33,34). It might be due to difference in socio demographic characteristics of the study participants in two study areas.

Contrary to study conducted in Cameroon and Boke Woreda, Ethiopia(18,36) parity was not shown to be linked with late ANC initiation. It might be due to the distribution of multipara and nulliparous among cases and controls were nearly similar and hidden the impacts of parity on late ANC follow up initiation. Pregnancy status was another insignificant variable in the current study. In contrast, the finding is different from studies conducted in Kembata and Gonder, Ethiopia(25,27) In the current study, the majority of respondents were married and probability of unplanned pregnancy is low. So that, fewer unplanned pregnancies among participants was the reason.

### Limitations

Misclassification bias might also be introduced since gestational age was determined using LMP based on the women’s memory.In addition, the current study focused solely on public health centers in Jimma town, with no mentions of public hospitals, private clinics, or private hospitals.

### Conclusion and recommendations

The study identified that, being age > 25 years, poor knowledge about ANC, previous home delivery, recognizing pregnancy through missed period, current pregnancy related pain before initiation, and lack of family support during ANC booking were determinants of late ANC initiation in the current study.Health care providers should provide health education at facility level regularly with due attention to late initiation of ANC, eligibility of all pregnant mothers regardless of feeling pregnancy related medical problems. While providing health education, special consideration should be given for mothers aged > 25 years.

## Data Availability

All data produced in the present study are available upon reasonable request to the authors

## Acknowledgment

We wouldlike to express our heartfelt appreciation to all participants of the study, Jimma University, and Jimma town health office for their unwavering support and dedication.

## Author Contributions

Conceptualization: Mohammed Jihad, KifleWoldemichael&YenealemGezehagn

Data curation: Mohammed Jihad.

Formal analysis: Mohammed Jihad

Funding acquisition: Mohammed Jihad.

Investigation: Mohammed Jihad, KifleWoldemichael&YenealemGezehagn

Methodology: Mohammed Jihad, KifleWoldemichael&YenealemGezehagn

Project administration: Mohammed Jihad.

Resources: Mohammed Jihad

Supervision: Mohammed Jihad, KifleWoldemichael&YenealemGezehagn

Visualization: Mohammed Jihad, KifleWoldemichael&YenealemGezehagn

Writing – original draft: Mohammed Jihad, KifleWoldemichael&YenealemGezehagn

Writing – review & editing: KifleWoldemichael&YenealemGezehagn

## Figures

<u>S1_Fig</u>. Proportional allocation of sample size among health centers in Jimma Town, 2021

<u>S2_Fig</u>. Knowledge status of the mothers among case and control groups, Jimma Town, 2021

## Supporting information

All data will be available from correspondence author based up on request.

