## Supporting information for "Determinants of Late Initiation for Antenatal Care Follow up Among Pregnant Mothers Attending Public Health Centers at Jimma Town, South West Ethiopia, 2021: Unmatched Case–Control Study"

### Questionnaire (English version)

| **Part I. Socio demographic questions** | | | |
| --- | --- | --- | --- |
| ***No.*** | ***Questions*** | ***Response*** | ***Remark*** |
| 101 | Age in years? | **____________** |  |
| 102 | Residence? | 1. Urban  2. Rural |  |
| 103 | Marital status? | 1. Single ( never married)  2. Married  3. Divorced/ separated  4. Widowed |  |
| 104 | Religion | 1. Muslim  2. Orthodox  3. Protestant  4. Catholic  5. Others [specify) |  |
| 105 | Family size | -------- |  |
| 106 | Educational status? | 1. Can’t read and write  2. primary (1-8)  3. secondary and above |  |
| 107 | Your occupational status | 1. Gov’t employed  2. Private employed  3. Merchant  4. Daily laborer  5. House wife  6. Other (specify ) ------- |  |
| 108 | Occupational status of your husband | 1. Gov’t Employed  2. Private employed  3. Merchant  4. Daily laborer  5. Farmer  6. Other |  |
| 109 | Educational status of your husband? | 1. Can’t read and write  2. primary (1-8)  3. secondary (9-12) and above |  |
| **Part II. Question on knowledge of ANC** | | | |
| 201 | What do you understand by antenatal care? | 1. It is regular medical and nursing care recommended for woman during pregnancy  2. It is to treat and prevent potential health problems throughout the course of the pregnancy.  3. It is to treat pregnant mothers with complication  4. Don’t know |  |
| 202 | Who are eligible women to visit health facility for ANC services? | 1. All pregnant women  2. Pregnant women with complication  3. I don’t know |  |
| 203 | What do you think would be the benefits of early ANC? | 1. For maternal health  2. For child health  3. For mother and child  4. I don’t know |  |
| 204 | At what weeks/gestational age do you think it is good to start ANC after amenorrhea? | 1. Before 16 weeks  2. 16-24 weeks  3. 28-36 weeks  4.When there is a problem/feeling ill  5. I don’t know |  |
| 205 | How many times do you think a woman needs to go for ANC at a health facility during pregnancy? | 1. Once during pregnancy  2. 2 times  3. 3 times  4. 4 times  5. More than 4 times  6. Others(specify)_______  7. I don’t know |  |
| **Part III. Maternal, medical and obstetrics factors related question** | | | |
| 301 | Have you experienced pregnancy before | 1. Yes  2. No | If no, skip to Q 306 |
| 302 | If yes Q.301, No. of gravida? | ----------- |  |
| 303 | If have any pregnancy experience for que. No. 301, is there any abortion? | 1.Yes  2.No |  |
| 304 | If you have any pregnancy experience before, for Q.No 301, have you had any ANC visit for last pregnancy? | 1.Yes  2.No |  |
| 305 | If yes for Q304, at what GA did you visit for first ANC service? | ------Weeks |  |
| 306 | Have you experienced birth before | 1. Yes  2. No | If no, skip to Q 312 |
| 307 | If yes Q.306, No. births | ------------ |  |
| 308 | If have any birth experience for que. No. 306, where did you deliver the last one? | 1.Health facility  2.Home |  |
| 309 | If you have any birth experience for que. No. 306, is there any still birth? | 1.Yes  2.No |  |
| 310 | If have any birth experience for que. No. 306, is there any CS history? | 1.Yes  2.No |  |
| 311 | If have any birth experience for que. No. 306, is there any neonatal death history? | 1.Yes  2.No |  |
| 312 | Is the current pregnancy planned? | 1.Yes  2.No | If no, skip to Q 314 |
| 313 | If yes for Q. No.311, does the plan include your spouse? | 1.Yes  2.No |  |
| 314 | At what GA/weeks did you start first ANC for current pregnancy? | --------Weeks |  |
| 315 | How did you know your pregnancy? | 1. By missed period  2. By Urine test |  |
| 316 | At what GA did you recognize your pregnancy? | ------ |  |
| 317 | Did you reveal your pregnancy immediately after you recognize? | 1Yes  2.No | If no, skip to Q 320 |
| 318 | If Yes for Ques 317, at what GA did you reveal? | ----- |  |
| 319 | To whom did you tell you become pregnant for the first time? | 1. Your Husband  2. Your Mother  3. Your Sister  4. Your Friend  5. Other (specify)_______________ |  |
| 320 | Is there any pregnancy related medical problem that makes you start ANC visit? | 1.Yes  2.No |  |
| **Part IV. Health service related questions** | | | |
| 401 | Distance from Health facility/ minutes it takes to reach? | _______________ |  |
| 402 | Is there any payment you have been asked for the service? | 1.Yes  2.No | If no, skip to Q 404 |
| 403 | Rate your satisfaction level on staff approach | 1. Highly satisfied  2. Satisfied  3. Medium  4. Dissatisfied  5.Highly Dissatisfied |  |
| 404 | Rate your satisfaction level on waiting time to get service | 1. Highly satisfied  2. Satisfied  3. Medium  4. Dissatisfied  5.Highly Dissatisfied |  |
| 405 | Rate your satisfaction level on privacy condition that is provided while check-up | 1. Highly satisfied  2. Satisfied  3. Medium  4. Dissatisfied  5.Highly Dissatisfied |  |
| 406 | Rate your satisfaction level on laboratory service | 1. Highly satisfied  2. Satisfied  3. Medium  4. Dissatisfied  5.Highly Dissatisfied |  |
| **Part V. Decision making and family support** | | | |
| 501 | With whom did you decide to initiate antenatal care visit for current pregnancy? | 1.With my husband  2. Myself  3. Others (specify) |  |
| 502 | With whom did you come to HC for ANC service for current pregnancy? | 1.With my husband  2. Myself  3. Others (Specify) |  |

**Part VI Wealth index Questionnaire**

| No | Items | Responses |
| --- | --- | --- |
|  |  | 1.Yes  2. No |
| **Land and Animals possession (Yes- 1 /No-2)** | | |
| 601 | Does your household own agricultural land? |  |
| 602 | Milk cows? |  |
| 603 | Goats? |  |
| 604 | Sheep? |  |
| 605 | Chickens |  |
| 606 | Donkey |  |
| **House equipment possession (Yes- 1 /No-2)** | | |
| 607 | Radio |  |
| 608 | Television |  |
| 609 | Bed |  |
| 610 | Mobile |  |
| 611 | Mattress |  |
| 612 | Refrigerator |  |
| 613 | Vehicles (Bajaj, Motor bicycle, Car…) |  |
| 614 | living house |  |
| 615 | Do you have a separate kitchen? |  |
| **Living house standard (circle the correct)** | | |

This is all what I want to ask you. Thank you for spending your time and

Valuable information you gave me. Do you have any question that I can address for you? _________________________________________________

Name of data collector ---------------- signatures---------

Name of supervisors------------------ signature------------
